# Large-Scale Hypothesis Testing for Causal Mediation Effects with Applications in Genome-wide Epigenetic Studies

**DOI:** 10.1101/2020.09.20.20198226

**Authors:** Zhonghua Liu, Jincheng Shen, Richard Barfield, Joel Schwartz, Andrea A. Baccarelli, Xihong Lin

## Abstract

In genome-wide epigenetic studies, it is of great scientific interest to assess whether the effect of an exposure on a clinical outcome is mediated through DNA methylations. However, statistical inference for causal mediation effects is challenged by the fact that one needs to test a large number of composite null hypotheses across the whole epigenome. Two popular tests, the Wald-type Sobel’s test and the joint significant test using the traditional null distribution are underpowered and thus can miss important scientific discoveries. In this paper, we show that the null distribution of Sobel’s test is not the standard normal distribution and the null distribution of the joint significant test is not uniform under the composite null of no mediation effect, especially in finite samples and under the singular point null case that the exposure has no effect on the mediator and the mediator has no effect on the outcome. Our results explain why these two tests are underpowered, and more importantly motivate us to develop a more powerful Divide-Aggregate Composite-null Test (DACT) for the composite null hypothesis of no mediation effect by leveraging epigenome-wide data. We adopted Efron’s empirical null framework for assessing statistical significance of the DACT test. We showed analytically that the proposed DACT method had improved power, and could well control type I error rate. Our extensive simulation studies showed that, in finite samples, the DACT method properly controlled the type I error rate and outperformed Sobel’s test and the joint significance test for detecting mediation effects. We applied the DACT method to the US Department of Veterans Affairs Normative Aging Study, an ongoing prospective cohort study which included men who were aged 21 to 80 years at entry. We identified multiple DNA methylation CpG sites that might mediate the effect of smoking on lung function with effect sizes ranging from −0.18 to − 0.79 and false discovery rate controlled at level 0.05, including the CpG sites in the genes AHRR and F2RL3. Our sensitivity analysis found small residual correlations (less than 0.01) of the error terms between the outcome and mediator regressions, suggesting that our results are robust to unmeasured confounding factors.

## 1 Introduction

Cigarette smoking is a well-known risk factor for reduced lung function (Tommola et al. 2016). It is thus of scientific interest to investigate the underlying causal mechanism and epigenetic pathway of the observed association between smoking and lung function. Motivated by the ongoing Normative Aging Genome-Wide Epigenetic Study that will be described in Section 6, we are interested in studying whether the effect of smoking on lung function is mediated by DNA methylations. DNA methylation is a heritable epigenetic mechanism that occurs by the covalent addition of a methyl (*CH*_3_) group to the base cytosine (C) at its 5-position within the CpG dinucleotide. The term CpG refers to the base cytosine (C) linked by a phosphate bond to the base guanine (G) in the DNA nucleotide sequence. Aberrations in the DNA methylations can affect downstream gene expressions and thus have an important role in the etiology of human diseases. There is increasing evidence that epigenetic mechanisms serve to integrate genetic and environmental causes of complex traits and diseases (Liu et al. 2013; Bind et al. 2014). Since DNA methylation is a reversible biological process (Wu and Zhang 2014), mediation analysis results can help discover novel epigenetic pathways as potential therapeutic targets.

Causal mediation analysis is a useful statistical method to answer the scientific question of whether DNA methylation mediates the effect of smoking on lung function. In the causal inference framework, the natural indirect effect (NIE) measures the evidence of mediation effect of an exposure on an outcome through a mediator (Robins and Greenland 1992; Pearl 2001) and is often of primary scientific interest. The classical regression approach to mediation analysis proposed by Baron and Kenny (1986) is a widely used method in social sciences for continuous outcomes and mediators, where the mediation effect is the product of the exposure-mediator and mediator-outcome effects, and is more generally referred to as the product method. This classical product method for mediation analysis is equivalent to the NIE defined in modern causal inference framework when the exposure-mediator interaction is absent (VanderWeele and Vansteelandt 2009; Valeri and VanderWeele 2013).

As the mediation effect is composed of the product of two parameters, MacKinnon et al. (2002) pointed out that the null hypothesis of no mediation effect is composite in the single mediation effect testing settings. Indeed, MacKinnon et al. (2002) found through simulation study that the Wald-type Sobel’s test (Sobel 1982) is overly conservative and thus underpowered, and recommended researchers to use the slightly more powerful joint significance test (also known as the MaxP test) for detecting mediation effects. However, both the Sobel’s test and the MaxP test perform poorly in genome-wide epigenetic studies as demonstrated empirically by Barfield et al. (2017). There are three reasons: (1) the association signals are generally weak and sparse with limited sample sizes; (2) the heavy multiple testing burden to be adjusted; (3) the composite null nature of the mediation effect testing that has not been taken into account.

For a variable to serve as a causal mediator in the pathway from an exposure to an outcome, it must satisfy the following two conditions simultaneously: (1) the exposure has an effect on the mediator; (2) the mediator has an effect on the outcome. The null hypothesis of no mediation effect is thus composite and consists of three cases: (1) the exposure has no effect on the mediator, the mediator has an effect on the outcome; (2) the exposure has an effect on the mediator, the mediator has no effect on the outcome; (3) the exposure has no effect on the mediator, and the mediator has no effect on the outcome. This salient feature of the composite null hypothesis imposes great statistical challenges for making inference on the mediation effect, and the uncertainty associated with the three cases under the composite null hypothesis should be taken into account when constructing valid and powerful testing procedures.

One attempt is the MT-Comp method proposed by Huang (2019), which however only works when the sample size is small, as the type I error rate of MT-Comp can be inflated when the sample size is large as stated in the original paper. This is because the MT-Comp method assumes that the association signals (an increasing function of the sample size) for the exposure-mediator or/and the mediator-outcome relationships are weak and sparse, which will be violated when the sample size is large. Therefore, it is pressing to develop statistically valid and powerful testing procedures to detect mediation effects that are suitable for general use in large-scale genome-wide epigenetic studies.

The main goal of this paper is to develop a valid and powerful large-scale testing procedure for detecting causal mediation effects by leveraging data from epigenome-wide DNA methylation studies. First, we study the statistical properties of the commonly used tests for causal mediation effects, Sobel’s test and the joint significance test. We show that the joint significance test is the likelihood ratio test for the composite null hypothesis of no mediation effect, and derive the null distributions of Sobel’s test and the joint significance test. Our results show that they follow non-standard distributions, and both the Sobel test and the joint significance test are conservative in the sense that their actual sizes are always smaller than the nominal significance level for any fixed sample size. The MaxP test is always more powerful than the Sobel’s test, but is still underpowered to detect mediation effects in genome-wide epigenetic studies. We also studied the powers of these two tests analytically and found that their powers are maximized when the association signals for the exposure-mediator and mediator-outcome relationships are of equal strength. Our results clearly and rigorously explain why these two popular tests are underpowered and thus are not suitable for large-scale inference for mediation effects.

To overcome the limitations of Sobel’s test and the joint significance test, we propose the Divide-Aggregate Composite-null Test (DACT), which improves the power by leveraging the whole genome DNA methylation data in a way that large-scale mediation effect testing is a blessing rather than a curse. Specifically, genome-wide data allow us to estimate the relative proportions of the three null cases that can be incorporated into the construction of the DACT test statistic as a composite *p*-value obtained by averaging the case-specific *p*-values weighted using the estimated case proportions. The DACT statistic follows a uniform distribution on the interval [0, 1] approximately if the exposure-mediator or the mediator-outcome association signals are sparse. It can depart from the uniform distribution when such signals are not sparse. To address this issue, we further propose to use Efron’s empirical null framework for inference (Efron 2004), where the empirical null distribution can be consistently estimated using the method developed by Jin and Cai (2007). We also study the statistical properties of the DACT method. We show that the proposed DACT method works well in both simulation studies and real data analysis of the Normative Aging Study (NAS), and outperforms Sobel’s test and the MaxP test substantially. We also perform a comprehensive sensitivity analysis to evaluate the robustness of our analysis results with respect to the no unmeasured confounding assumption.

The rest of our paper is organized as follows. In Section 2, we present the regression models for causal mediation analysis, derive the null distributions of Sobel’s test and the MaxP test, and then discuss the limitations of these two tests in genome-wide epigenetic studies. In Section 3, we propose the DACT testing procedure and study its statistical properties. In Section 4, we discuss the connections and differences of Sobel’s test, the MaxP test and our DACT. In Section 5, we conduct extensive simulation studies to evaluate the type I error rates of DACT along with Sobel’s test and the joint significant test, and compare their powers under various alternatives. In Section 6, we apply the DACT method to the Normative Aging Genome-Wide Epigenetic Study to detect the mediation effects of DNA methylation CpG sites in the causal pathway from smoking behavior to lung function. The paper ends with discussions in Section 7.

## 2 Causal Mediation Analysis

### 2.1 Assumptions and Regression Models

Let *A* denote an exposure, *Y* a continuous outcome, *M* a continuous mediator and ***X*** additional covariates to adjust for confounding. Baron and Kenny (1986) proposed the following linear structural equation models for the outcome and the mediator

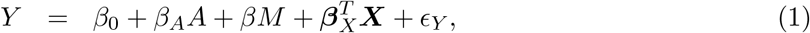

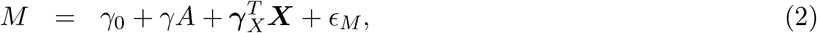

where *ϵ*_*Y*_ and *ϵ*_*M*_ are the error terms with mean zeros and constant variances, which are also uncorrelated under the standard assumptions (1)-(5) stated below in causal mediation analysis (Imai et al. 2010). The constant variance assumption was found to be reasonable when the methylation level is on the M-value scale (Du et al. 2010). It is well-known that the least squares estimation method gives unbiased parameter estimators in models (1) and (2). If the outcome *Y* is binary and rare, then we can fit the following logistic models using the maximum likelihood estimation (MLE) method

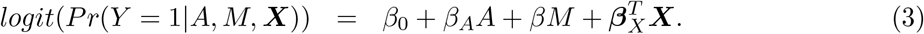

Our primary interest is the so called Natural Indirect Effect (NIE) defined by Robins and Greenland (1992) and Pearl (2001), which measures the effect of the exposure on the outcome mediated through the mediator. In the modern causal inference framework, one assumes the following standard identification assumptions for estimating the NIE (VanderWeele and Vansteelandt 2009; Valeri and VanderWeele 2013): (1) There are no unmeasured exposure-outcome confounders given ***X***; (2) There are no unmeasured mediator-outcome confounders given (***X***, *A*); (3) There are no unmeasured exposure-mediator confounders given ***X***; (4) There is no effect of exposure that confounds the mediator-outcome relationship; (5) There is no exposure and mediator interaction on the outcome. Under these standard assumptions, the NIE (mediation effect) can be identified. When both the mediator and the outcome are continuous, the NIE is equal to *βγ*. When the mediator is continuous, and the outcome is binary and rare, the NIE is approximately equal to *βγ* on the log-odds-ratio scale (Valeri and VanderWeele 2013). Graphically, the NIE measures the effect of the causal chain *A* → *M* → *Y* as shown in a directed acyclic graph (DAG) in Figure 1. We assume that the covariates (possibly vector-valued) ***X*** contain all the confounders. In practice, there might be unmeasured confounders *U* omitted from mediation analysis. Sensitivity analysis can be performed to assess the robustness of data analysis results, for example, using the method proposed by Imai et al. (2010) as we will do in Section 6.

**Figure 1:**
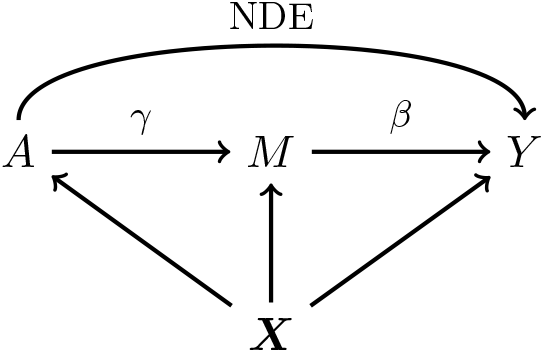
A causal DAG for mediation analysis. *A* is the exposure, *M* is the mediator, *Y* is the outcome and ***X*** represents measured confounders. *γ* is the causal effect of *A* on *M* and *β* is the causal effect of *M* on *Y*. NDE stands for Natural Direct Effect.

Under the assumptions (1)-(5), the causal effect of the exposure *A* on the mediator *M* is orthogonal to the causal effect of the mediator *M* on the outcome *Y* (Figure 1). We now show this simple but important result, which will be used in Section 3 to simplify the estimation and testing procedure. Specifically, under the assumptions (1)-(5), the joint probability density function of (*Y, M, A*, ***X***) can be factored as *f* (*Y, M, A*, ***X***; **Θ**) = *f* (*Y* |*M, A*, ***X***; **Θ**_1_)*f* (*M*|*A*, ***X***; **Θ**_2_)*f* (*A*, ***X***), where *f* (*A*, ***X***) can be discarded because it is ancillary for the model parameters in equations (1) - (3). Therefore, we only need *f* (*Y* |*M, A*, ***X***; **Θ**_1_)*f* (*M*|*A*, ***X***; **Θ**_2_) for the inference of unknown parameters in the models (1) - (3). The *A* → *M* association captured by *γ* is contained in *f* (*M*|*A*, ***X***; **Θ**_2_), and the *M* → *Y* association captured by *β* is contained in *f* (*Y* |*M, A*, ***X***; **Θ**_1_). Denote the log-likelihood as *𝓁*(*·*), then we have *∂𝓁*(*·*)*/∂β∂γ* = 0 as *β* only appears in the **Θ**_1_ and *γ* only appears in the **Θ**_2_. This implies that the two parameters *β* and *γ* are orthogonal. A more detailed proof is provided in the Supplementary Materials. We will use this result throughout the whole paper.

In genome-wide epigenetic studies, we are interested in assessing whether a particular DNA methylation CpG site lies in the causal pathway from an exposure to a clinical outcome. This can be formulated as the following hypothesis testing problem

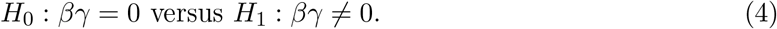

As mentioned in Section 1, the null hypothesis *H*_0_: *βγ* = 0 is composite and the null parameter space can be decomposed into three disjoint cases,

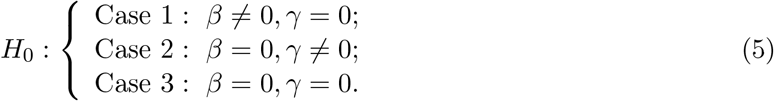

The fourth case: *β≠* 0, *γ≠* 0 corresponds to the alternative hypothesis. In practice, we can fit the outcome and mediator regression models and obtain consistent estimates 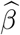 and 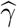 for the regression coefficients *β* and *γ* respectively. We have the following standard normal approximation

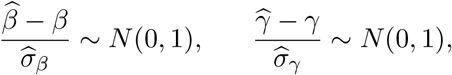

where 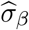 and 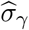 are the estimated standard errors for 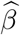 and 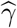 respectively. A consistent point estimator for the mediation effect is 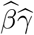. A rejection of the null hypothesis *H*_0_: *βγ* = 0 suggests the presence of a mediation effect by *M*.

### 2.2 The Wald-type Sobel’s Test

Using the first-order multivariate delta method, Sobel (1982) obtained the standard error for the product-method estimator 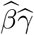 and proposed the following test statistic to detect the mediation effect

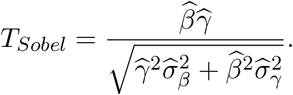

Note that the covariance term between 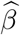 and 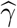 was set to zero here because 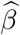 and 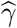 are independent of each other. To determine statistical significance, Sobel (1982) used the standard normal distribution as the reference distribution to calculate the *p*-value of *T*_*Sobel*_. MacKinnon et al. (1998) found that the Sobel’s test has low power via simulation studies but did not explain theoretically why the Sobel’s test is underpowered.

To provide statistically rigorous guidance for applied researchers when using Sobel’ test, we now investigate the statistical properties of Sobel’s test and show why it is underpowered. First, we show that under the composite null, Sobel’s test is conservative for any finite sample size but has correct type I error rate asymptotically in the null Case 1 and Case 2. While in the null Case 3, Sobel’s test is always conservative even asymptotically. The fundamental reason is that the first-order multivariate delta method fails because the gradient is (0, 0), and the usual asymptotic normal approximation for the null distribution of Sobel’s test is thus incorrect in the null Case 3. Our result explains clearly and rigorously why Sobel’s test is underpowered.

For the ease of exposition, we introduce some notation. Denote 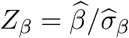 and 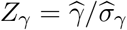. We write *Z*_*β*_ as *Z*_*β*_ (*n*) and *Z*_*γ*_ as *Z*_*γ*_ (*n*) to emphasize that those two statistics depend on the sample size *n*. Direct calculation gives

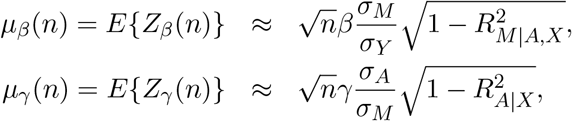

where *σ*_*A*_ is the coefficient of determination by is the standard deviation of exposure *A*, 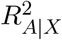 regressing exposure *A* on the covariates *X*, and 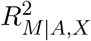 is the coefficient of determination by is the coefficient of determination of the mediator regression model (2). In what follows, *µ*_*γ*_ (*n*) and *µ*_*β*_ (*n*) will be referred to as the association signals for the exposure-mediator and mediator-outcome relationships respectively.

It is reasonable to assume that 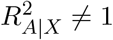 and 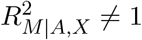. We then can rewrite *T*_*Sobel*_ as

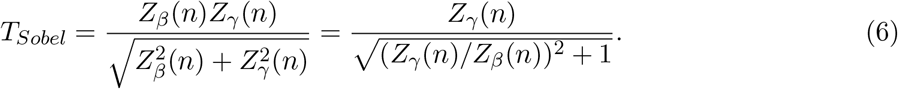

This representation of Sobel’s test statistic can help us better understand its behavior. In the null Case 1, the size of Sobel’s test is strictly smaller than the nominal significance level *α* for any finite sample size by noting the following result

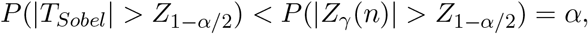

where *Z*_1−*α/*2_ denotes the 1 − *α/*2 percentile of the standard normal distribution. We observe that the conservativeness of Sobel’s test in null Case 1 can be alleviated when the sample size goes to infinity. To show this result, without loss of generality, we can assume that *β >* 0. Then we have *µ*_*β*_ (*n*) → +*∞* as the sample size *n* → *∞*. It can be easily seen that {*Z*_*β*_ (*n*)}^−1^ converges to zero and *Z*_*γ*_ (*n*) is bounded in probability, therefore the ratio *Z*_*γ*_ (*n*)*/Z*_*β*_ (*n*) converges to zero in probability. Using Slutsky’s theorem, *T*_*Sobel*_ follows the standard normal distribution asymptotically. Therefore, the Sobel’s test has correct size asymptotically, but is conservative for finite sample sizes in Case 1. The same conclusion holds in the null Case 2.

In the null Case 3, the ratio *Z*_*γ*_ (*n*)*/Z*_*β*_ (*n*) is stochastically bounded and in fact follows the standard Cauchy distribution asymptotically. The central limit theorem cannot be applied to the test statistic *T*_*Sobel*_ in this case and the asymptotic distribution of *T*_*Sobel*_ is not the standard normal, but is normal with mean zero and variance equal to 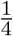 asymptotically. This explains why it is incorrect to use the standard normal distribution as the reference distribution to calculate *p*-value for Sobel’s test. The actual type I error rate is much smaller than the nominal significance level *α* even asymptotically. The conservativeness of Sobel’s test cannot be alleviated in the Case 3 even with increased sample size. We summarize our findings about Sobel’s test in Result 1, with proofs provided in the Supplementary Materials.

**Result 1** *Sobel’s statistic T*_*Sobel*_ *for testing the composite null of no mediation effect (4) has the following properties:*

a. 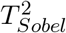 *follows the same distribution as the inverse of the sum of two independent standard Lévy variables (inverse chi-squared random variables with one degree of freedom) asymptotically*.
b. *Under the composite null (4), in Cases 1 and 2, T*_*Sobel*_ *follows N* (0, 1) *asymptotically; In Case 3, T*_*Sobel*_ *follows* 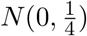 *or equivalently* 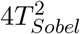 *follows* 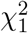 *distribution asymptotically*.
c. *The power of the Sobel test given the significance level α can be calculated analytically as*

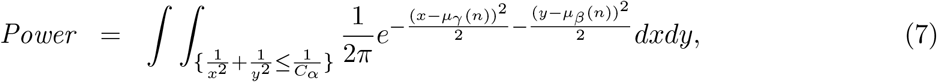 *where C*_*α*_ *is the critical value at the significance level α. The power of the Sobel’s test is maximized when* |*µ*_*β*_ (*n*)| = |*µ*_*γ*_ (*n*)| *for a fixed NIE signal strength*.

Figure 2 shows the empirical distributions of *T*_*Sobel*_ in the null Case 1 (upper panels) and Case 3 (lower panels). In the null Case 1, we set *β* = 0.2, *γ* = 0; while in the null Case 3, we set *β* = *γ* = 0. Sample sizes *n* = 100, 500, 5000 are considered in both Case 1 and Case 3. We first generate random samples for *Z*_*β*_ and *Z*_*γ*_, and then use the formula (6) to get random samples for *T*_*Sobel*_. The density function of *T*_*Sobel*_ was estimated by the kernel density estimator using the R function *density* with its default setting. We then compare the density function plots of *T*_*Sobel*_ to the standard normal density function under various scenarios. We found that the normal approximation for the test statistic *T*_*Sobel*_ improves as the sample size increases in the null Case 1, but the standard normal approximation fails even with increased sample size in the null Case 3.

**Figure 2:**
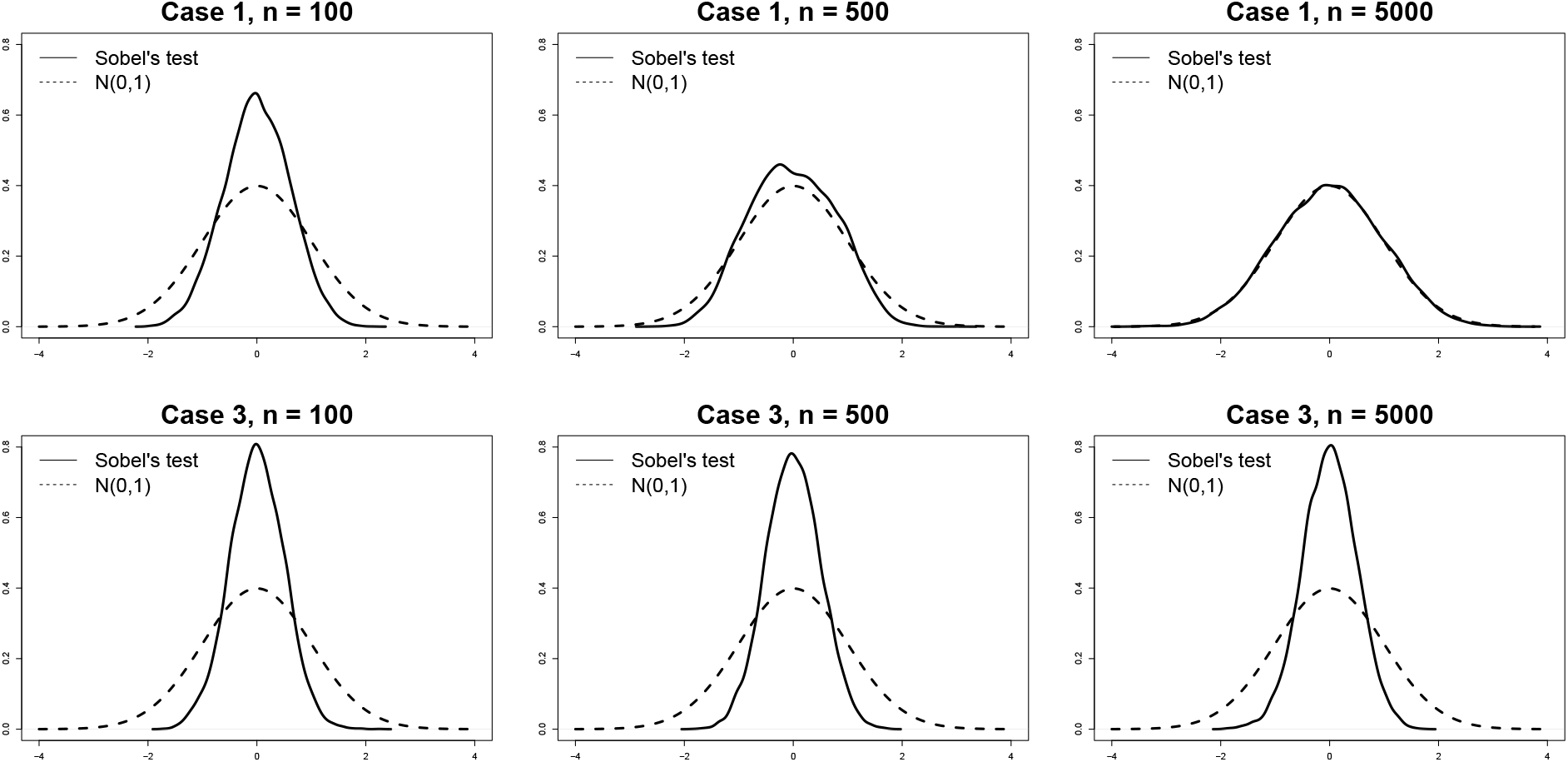
The kernel density estimates (the solid lines) of the probability density functions of *T*_*Sobel*_ in the null Case 1 and Case 3 with increasing sample sizes. 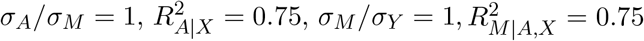. The upper panel is for null Case 1 (*β* = 0.2, *γ* = 0) and the lower panel is for null Case 3 (*β* = *γ* = 0) with sample sizes *n* = 100, 500, 5000. In null Case 3, the variance of *T*_*Sobel*_ is estimated to be 0.25. The dashed lines represent the probability density functions of the standard normal *N* (0, 1).

### 2.3 The Joint Significance (MaxP) Test

The joint significance test, also known as the MaxP test (MacKinnon et al. 2002), was developed based on the argument we have already stated in Section 1 that one can claim the presence of mediation effects if the following two c onditions a re s atisfied si multaneously: (1) the exposure has an effect on the mediator; (2) the mediator has an effect on the outcome. Let *p*_*β*_ = 2(1 − Φ(|*Z*_*β*_ |)) be the *p*-value for testing *H*_0_: *β* = 0, which is uniformly distributed on the interval [0, 1] when *β* = 0 holds, and will converge to zero in probability when *β ≠* 0. Let *p*_*γ*_ = 2(1 − Φ(|*Z*_*γ*_|)) be the *p*-value for testing *H*_0_: *γ* = 0, which is uniformly distributed on the interval [0, 1] when *γ* = 0 holds, and will converge to zero in probability when *γ* ≠ 0. Define MaxP = max(*p*_*β*_, *p*_*γ*_). Then, the MaxP test declares statistical significance for testing the composite null *H* _0_: *β γ* = 0 if MaxP *< α*. Intuitively, the MaxP test requires that both *p*_*β*_ and *p*_*γ*_ are significant by rejecting b oth *H* _0_: *β* = 0 and *H*_0_: *γ* = 0 individually. This testing procedure has an intuitive appeal and is easy to interpret, and hence has been widely used by applied researchers. MacKinnon et al. (2002) found that the MaxP test is slightly more powerful than Sobel’s test using simulation studies, but did not provide any theoretical explanation for this empirical observation.

We now show that the MaxP test is conservative for testing *H*_0_: *βγ* = 0 in all the three null cases for any finite sample size. First, since *p*_*β*_ and *p*_*γ*_ are independent, we have

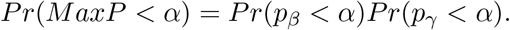

In Case 1, *Pr*(*p*_*γ*_ *< α*|*γ* = 0) = *α* and *Pr*(*p*_*β*_ *< α*|*β ≠* 0) *<* 1 for any finite sample size, so *Pr*(*MaxP < α*) *< α*. Thus, the MaxP test is conservative in Case 1 for any finite sample size. However, if the sample size goes to infinity, then

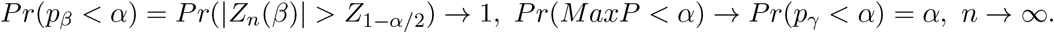

Therefore, the MaxP test has correct size and is equivalent to *p*_*γ*_ asymptotically in the null Case 1. Likewise, the MaxP test has correct size and is equivalent to *p*_*β*_ asymptotically in the null Case 2. In the null Case 3, both *p*_*β*_ and *p*_*γ*_ are uniformly distributed. Therefore, *Pr*(*p*_*β*_ *< α*) = *Pr*(*p*_*γ*_ *< α*) = *α*, and *Pr*(*MaxP < α*) = *α*^2^ *< α* for any *α* ∈ (0, 1) and any sample size. Thus, the MaxP test is always conservative regardless of sample size in the null Case 3. Traditionally, the MaxP test statistic itself is treated as a *p*-value, which is correct in the null Case 1 and 2 asymptotically, but is incorrect in the null Case 3. In the next section, we will propose anew testing procedure that can greatly improve the power of the MaxP test in large-scale multiple testing settings.

Result 2 states that the MaxP test is the likelihood ratio test (LRT) for the composite null *H*_0_: *βγ* = 0 and the power of the MaxP test can also be calculated analytically. Its proof is given in the Supplemental Materials.

**Result 2** *The joint significance test (MaxP test) has the following properties:*

a. *The MaxP test is the likelihood ratio test for the composite null of no mediation effect*.
b. *The exact cumulative distribution function of the MaxP statistic in Case 1 is*

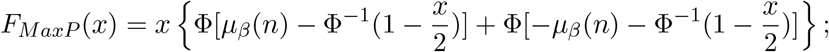 *and similarly for Case 2 by changing µ*_*β*_ (*n*) *to µ*_*γ*_ (*n*); *The MaxP statistic follows Beta*(2, 1) *distribution in Case 3, and the uniform distribution on [0,1] asymptotically in Case 1 and 2*.
c. *The power of the MaxP test given the significance level α can be calculated analytically as*

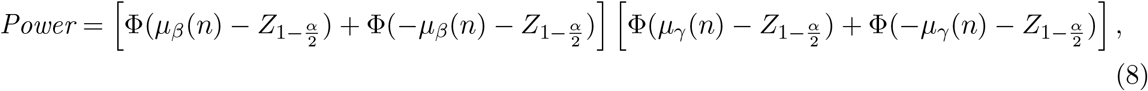 *and the power of MaxP test is maximized when* |*µ*_*β*_ (*n*)| = |*µ*_*γ*_ (*n*)| *for a fixed NIE strength*.

## 3 The Divide-Aggregate Composite-null Test (DACT)

### 3.1 Estimation of the Proportions of the Three Null Cases

In view of the conservativeness of Sobel’s test and the MaxP test, we propose in this section the Divide-Aggregate Composite-null Test (DACT) by leveraging information across a large number of tests in genome-wide epigenetic studies. Suppose that an Oracle knows the true relative proportions of the three null cases, then such information can be incorporated to increase the power of the MaxP test. A single test for mediation effects using either Sobel’s test or the MaxP test is challenged by the fact that one does not know which of the three null cases holds. Fortunately, we can obtain such information in modern large-scale multiple testing settings, such as in genome-wide epigenetic studies, where we can estimate the relative proportions of the three null cases based on a large number of tests across the whole genome. It is thus one of the few instances where high-dimensionality is not a curse but rather a blessing if used properly.

Suppose that there are a total of *m* DNA methylation CpG sites, where *m* is in the order of hundreds of thousands. For example, there are 484,613 CpG sites in the NAS data set to be described in details later in Section 6. To identify putative CpG sites lying in the causal pathway from the exposure to the outcome of interest, we need to perform a total of *m* hypothesis tests to assess the strength of the evidence against the composite null hypothesis *H*_0_: *βγ* = 0. There are *m* null hypotheses for the parameter *β* in the outcome regression model: 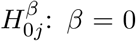, and *m* null hypotheses for the parameter *γ* in the mediator regression model: 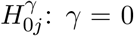, where 1 ≤ *j* ≤ *m*. We now define 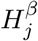 (the same for 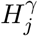) as a sequence of (possibly dependent) Bernoulli random variables, where 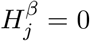 if 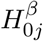 is true and 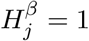 if 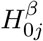 is false, 1 ≤ *j* ≤ *m*, a framework proposed by Efron et al. (2001) and later adopted widely (Storey 2002; Genovese and Wasserman 2004).

As shown in Section 2 that *β* and *γ* are orthogonal, we have that 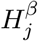 is independent of 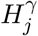 for 1 ≤ *j* ≤ *m*. For each DNA methylation CpG site, we fit the outcome and mediator regression models to obtain *p*-values *p*_*βj*_ for testing *β* and the *p*-values *p*_*γj*_ for testing *γ*, where 1 ≤ *j* ≤ *m*. Following Efron et al. (2001), assume that 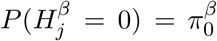 and 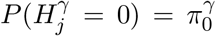, where the parameters 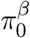 is the proportion of CpG sites that are not associated with the outcome under the outcome models (1) or (3), and 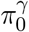 is the proportion of CpG sites that are not associated with the exposure in the mediator model (2). Since 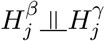, 1 ≤ *j* ≤ *m*, then we have

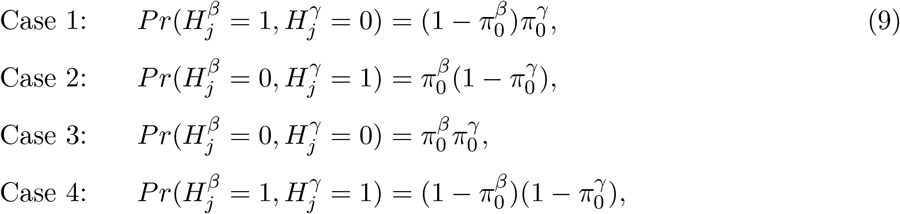

where Cases 1-3 together constitute the composite null hypothesis of null mediation effects, and Case 4 represents the alternative of non-null mediation effects.

Under the composite null *H*_0_: *βγ* = 0, the normalized relative proportions of the three null cases *w*_1_, *w*_2_, *w*_3_ are: 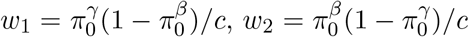 and 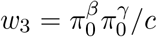 respectively, where the normalizing constant 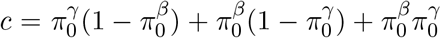, and *w*_1_ + *w*_2_ + *w*_3_ = 1. In typical epignome-wide association studies (EWAS), both 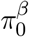 and 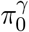 are close to one as in our NAS data set in Section 6. To be more general, we do not impose such sparsity assumption on our method.

We used the method proposed by Jin and Cai (2007), which is referred to as the JC method hereafter, to estimate 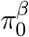 and 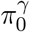 based on the *z*-scores for testing *β* = 0 and the *z*-scores for testing *γ* = 0 respectively. Suppose that we have *m* test statistics *z*-scores 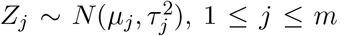, 1 ≤ *j* ≤ *m*, where *µ*_*j*_ = *µ*_0_ and 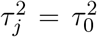 under the null. Here, we can set *µ*_0_ = 0 and *τ*_0_ = 1. Jin and Cai(2007) proposed to use the empirical characteristic function and Fourier analysis for estimating the proportion of true nulls. The empirical characteristic function is

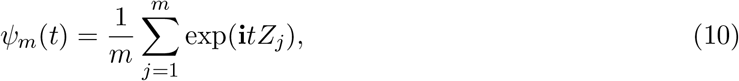

where 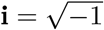. For *r* ∈ (0, 1*/*2), the proportion of true nulls *π*_0_ can be consistently estimated as

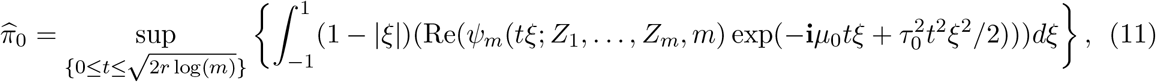

where Re(*x*) denotes the real part of the complex number *x*. Jin and Cai (2007) showed that 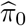 is uniformly consistent over a wide class of parameters for independent and dependent data under regularity conditions.

Kang (2020) also found in a recent simulation study that the JC method outperforms other competitors under practical dependence structures in genomic data. Here, we employ the JC method to estimate 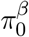 and 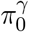 separately, and obtain uniformly consistent estimators 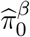 and 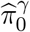.Then *w*_1_, *w*_2_, *w*_3_ are estimated by plugging in 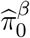 and 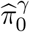 for the unknown parameters 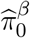 and 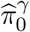 respectively. It is straightforward to show the resulting estimators ŵ_1_, ŵ_2_, ŵ_3_ are also consistent under the same regularity conditions of Jin and Cai (2007) using the continuous mapping theorem (van der Vaart 2000, pp. 7).

### 3.2 Construction of the Divide-Aggregate Composite-null Test (DACT)

We propose in this section the Divide-Aggregate Composite-null Test (DACT) for the composite null of no mediation effect *H*_0_: *βγ* = 0. We first consider how to perform mediation effect testing in each of the three null cases as defined in Section 2. In the null Case 1: *β≠* 0, *γ* = 0, we only need to test whether *γ* = 0 using the *p*-value *p*_*γ*_ because *β≠* 0. Similarly, in the null Case 2: *β* = 0, *γ≠* 0, we only need to test whether *β* = 0 using the *p*-value *p*_*β*_ because *γ≠* 0. While in the null Case 3: *β* = 0, *γ* = 0, we need to test whether both *β* and *γ* are nonzero. We can reject the null Case 3 if max(*p*_*γ*_, *p*_*β*_) *< α* at the significance level *α*. Intuitively, this requires that both *p*_*β*_ and *p*_*γ*_ are statistically significant. The *p*-value of the MaxP test can be computed as (MaxP)^2^ by noting that the MaxP test follows *Beta*(2, 1) distribution in the null Case 3 as given in the Result Following this logic, we propose the following case-specific *p*-values for testing mediation effects for the *j*th CpG site as

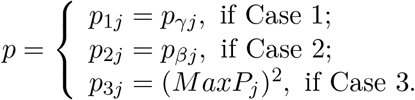

We now construct the DACT statstistic to test for the composite null of no mediation effect *H*_0_: *βγ* = 0 by using a composite *p*-value as a test statistic, which is calculated as follows:

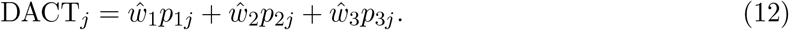

If any of *w*_1_, *w*_2_ and *w*_3_ is close to one, then the DACT statistic follows the uniform distribution on the interval [0, 1] approximately. Based on our empirical observation from the NAS data analysis in Section 6, *w*_3_ is very close to one. However, there are also scenarios when investigators want to conduct a more focused study within a smaller set of epigenetic markers from pre-screening studies, or based on prior knowledge (Cecil et al. 2014). In such circumstances, *w*_1_ or *w*_2_ may be anon-ignorable percentage, and the DACT statistic may depart from the uniform distribution on the interval [0, 1]. To make the DACT method applicable to those settings, we need to estimate the empirical null distribution of DACT.

We adopt Efron’s empirical null inference framework (Efron 2004) to calibrate the *p*-values of the DACT statistics by accounting for possible correlations among the tests. Specifically, we transform the DACT statistic using the inverse normal cumulative distribution function (CDF)

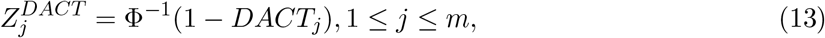

where Φ(*·*) denotes the standard normal CDF. Those *m* test statistics fall into two categories: 1) null mediation effects; 2) non-null mediation effects. Therefore, the marginal probability density function of 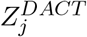 is

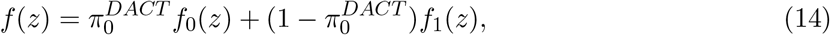

where 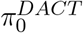 denotes the proportion of null mediation effects, *f*_0_(*z*) denotes the null distribution *N* (*δ, σ*^2^) and *f*_1_(*z*) denotes the non-null distribution.

Our goal here is to estimate *f*_0_(*z*) by estimating *δ* and *σ*^2^. The empirical characteristic function of 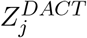 is 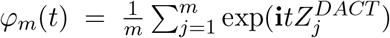. The expected characteristic function is 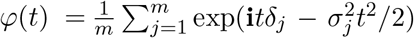, which can be decomposed as 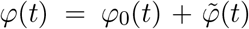, where 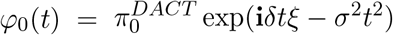 and 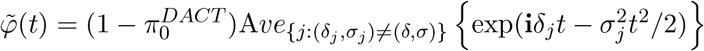.

Jin and Cai (2007) showed that for all *t* ≠ 0,

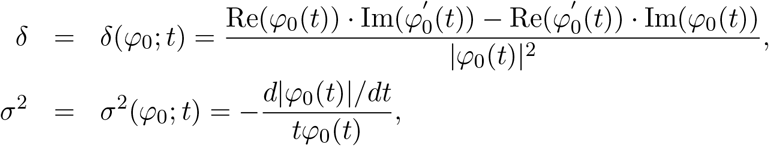

where Re(*x*), Im(*x*) and |*x*| denote the real part, the imaginary part and the modulus of the complex number *x*. For an appropriately chosen large *t, φ*_*m*_(*t*) ≈ *φ*(*t*) ≈ *φ*_0_(*t*), so that the contribution of non-null mediation effects to the empirical characteristic function is negligible. In practice, *t* is chosen as 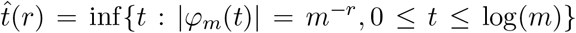, for a given *r* ∈ (0, 1*/*2). One then estimates *δ* and *σ*^2^ using

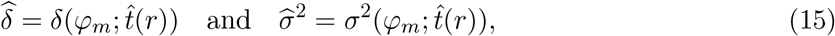

with *r* = 0.1 as recommended by Jin and Cai (2007). The two estimators 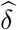 and 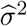 have been shown to be uniformly consistent for independent and dependent data under some regularity conditions (Jin and Cai 2007), and hence the empirical null probability density function estimator 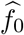 and the corresponding CDF estimator 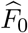 are both consistent. We then calibrate the *p*-value of 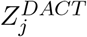 by

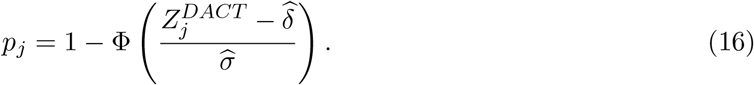

Efron’s empirical null framework is really a statement about the nature or the choice of the null distribution, and does not depend on the inference method to be used later for thresholding the test statistics (Schwartzman et al. 2009). If the empirical null is *N* (*δ, σ*^2^), then any method for controlling family-wise error rate (FWER) can be applied to the normalized *z*-scores *Z*^*\**^ = (*Z*−*δ*)*/σ* or equivalently the calibrated *p*-values. The FWER is controlled asymptotically as long as the empirical null distribution can be consistently estimated. The proof is trivial and thus omitted. The same argument also applies to the local and tail area false discovery rate (FDR) control (Efron et al. 2001; Efron 2004, 2010). The local FDR is defined as 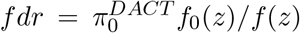 and the tail area FDR is 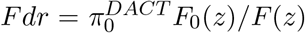, where *F*_0_(*z*) and *F* (*z*) are the corresponding CDFs of *f*_0_(*z*) and *f* (*z*) respectively. The parameter 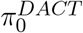 can be consistently estimated using the generic formula (11) by replacing *µ*_0_, *τ*_0_, *Z*_*j*_ by 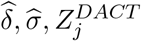 respectively. The marginal probability density function *f* (*z*) can be consistently estimated using the kernel density estimator 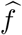 (Wasserman 2006, pp. 133), and the marginal CDF *F* (*z*) can be consistently estimated using the empirical CDF 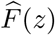 according to the classical Glivenko–Cantelli theorem (van der Vaart 2000, pp. 266). We show in the Supplemental Materials that the (local) FDR can be controlled asymptotically. We summarize our findings about DACT in Result 3.

**Result 3** *The proposed DACT has the following properties:*

a. *In Case 1 or Case 2, the DACT is asymptotically equivalent to both the Sobel’s test and the MaxP test*.
b. *In Case 3, the DACT has the correct size, while both Sobel’s test and the MaxP test are conservative for any sample size*.
c. *Under regularity conditions of Jin and Cai (2007)*, 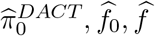 *are consistent estima-tors of e*_0_, *f*_0_, *f respectively. The local FDR for the jth composite null test* 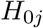 *is estimated as* 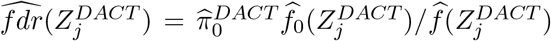. *Then the following procedure controls local FDR asymptotically at a pre-specified level q* ∈ [0, 1],

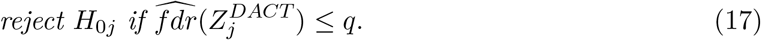 *The same result holds for the tail-area FDR control by replacing* 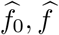 *by* 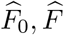 *respectively*.

#### Remark

The use of the empirical null distribution to correct bias and inflation of the observed *p*-values has been proven useful and effective in epigenome-wide association studies(van Iterson et al. 2017). If the genomic inflation factor *λ* of DACT is close to one, then this correction makes little change. However, if none of the three null cases is close to one, for example, when *w*_1_ = *w*_2_ = *w*_3_ = 1*/*3 as shown in Figure 4, then the corrected DACT (calibrated *p*-value for DACT) using equation (16) performs much better as demonstrated in our simulation studies in Section 5.

## 4 Comparison of the Three Tests

Our proposed data-adaptive DACT approach leverages information contained in the whole epigenome, and thus improves the power for testing mediation effects. Figure 3 shows that the rejection region of the MaxP test is a subset of the rejection region of the proposed DACT method, while the rejection region of Sobel’s test is a subset of the rejection region of the MaxP test. In other words, the DACT test dominates the MaxP test, and the MaxP test dominates Sobel’s test. Moreover, the rejection region of the DACT test depends on the relative proportions of the three null cases, as shown in Figure 3. Specifically, the DACT test has a larger rejection region in the presence of a larger proportion of Case 3.

**Figure 3:**
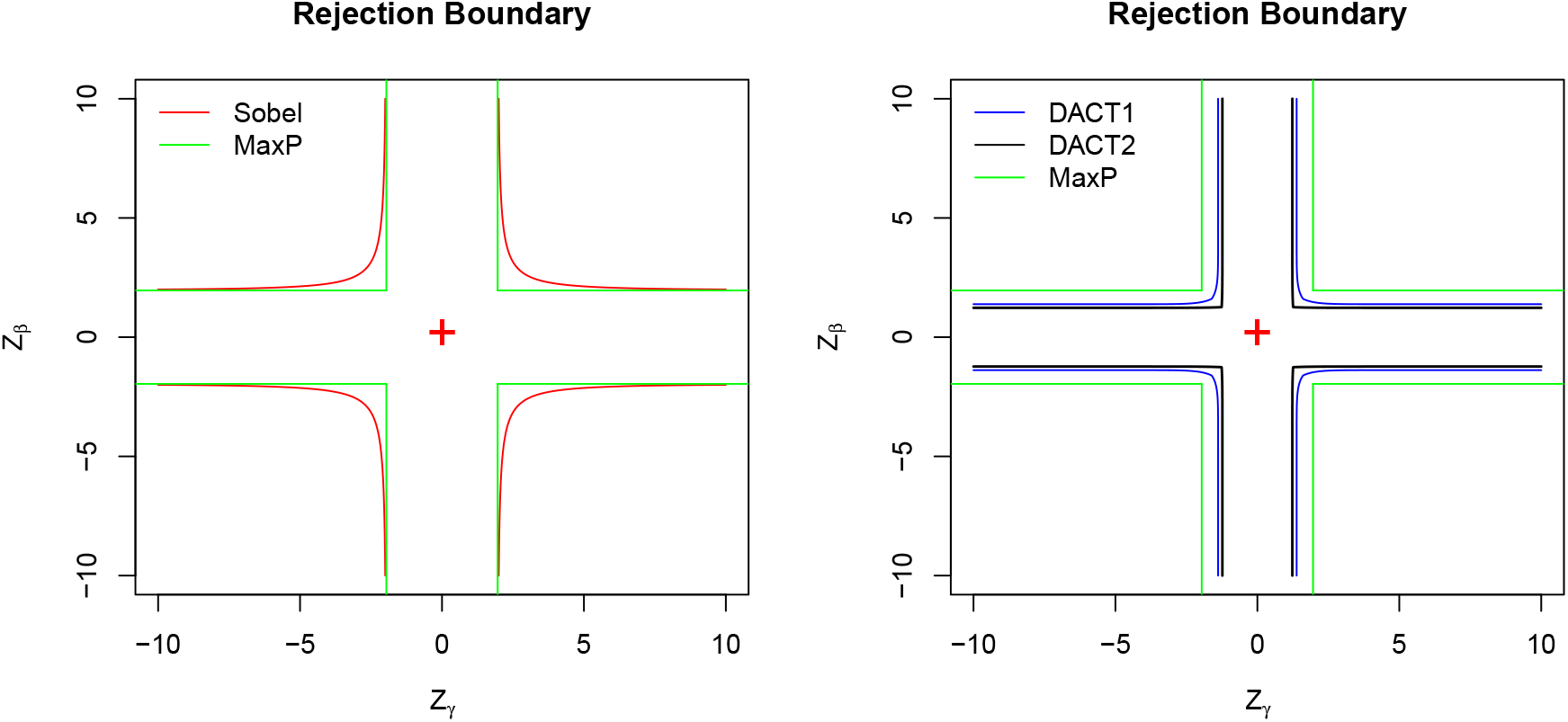
The rejection boundaries of the Sobel’s test, the MaxP test and the DACT test are plotted at significance level 0.05 on the *z*-score scale. For DACT, we consider two settings: setting 1 (denoted as DACT1): *w*_1_ = *w*_2_ = 0.2 and *w*_3_ = 0.6; setting 2 (denoted as DACT2): *w*_1_ = *w*_2_ = 0.02 and *w*_3_ = 0.96.

Formally, let’s compare the Sobel’s test and the MaxP test in finite sample settings. We already know that |*T*_*Sobel*_| *<* min(|*Z*_*β*_|, |*Z*_*γ*_|), therefore we have

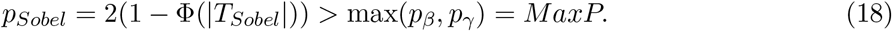

This result says that the MaxP test is always more significant than Sobel’s test at the significance level *α*. In other words, if the Sobel’s test detects a mediation effect, then the MaxP test will do as well, but not vice versa. Therefore, the MaxP test is uniformly more powerful than the Sobel’s test for any given significance level. In this regard, the Sobel’s test is inadmissible. However, the Sobel’s test and the MaxP test are asymptotically equivalent in Case 1 and 2. In Case 1, because *T*_*Sobel*_ is asymptotically equivalent to *Z*_*γ*_ and MaxP is asymptotically equivalent to *p*_*γ*_, therefore the inference using *T*_*Sobel*_ is asymptotically equivalent to MaxP. The same conclusion holds in Case 2 as well. In Case 3, the inferences using *T*_*Sobel*_ and MaxP are asymptotically different. The asymptotic *p*-value of *T*_*Sobel*_ is calculated using the normal distribution *N* (0, 1*/*4), while the asymptotic *p*-value of the MaxP test is calculated using the Beta distribution *Beta*(2, 1).

One can also show that the MaxP test based on *MaxP* = max(*p*_*β*_, *p*_*γ*_) can be equivalently defined using 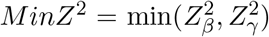. Both give the same inference. This provides a more clearer relationship of Sobel’s test and the MaxP test on the same scale directly using *Z*_*β*_ and *Z*_*γ*_. Specif-ically, since 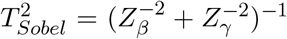, both 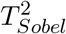 and *MinZ*^2^ asymptotically follow 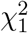in Cases 1 and 2. However, in Case 3, 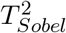 asymptotically follows 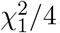, while *MinZ*^2^ asymptotically follows the distribution of the first order statistic of two independent random variables that follow the 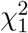 distribution, i.e., the distribution of 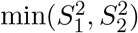, where 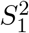 and 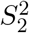 are independent random vari-ables that follow the 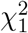 distribution. In Case 3, it is straightforward to show that the cumulative distribution function of *MinZ*^2^ is

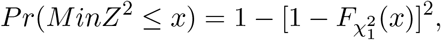

where 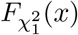 denotes the cumulative distribution function of a central chi-squared random variable with one degree of freedom. Therefore, in Case 3, the Wald-type Sobel’s test and the likelihood ratio test equivalent MaxP test have different distributions in both finite and large sample settings. In Section 2, we have shown that the actual sizes of the Sobel’s test and the MaxP test are smaller than the pre-specified nominal type I error rate *α*. Those two tests are thus underpowered because they do not fully spend the allowed amount of type I error *α*.

## 5 Simulation Studies

### 5.1 Type I Error Rates

In this section, we conduct extensive simulation studies to evaluate the type I error rate of the DACT method under the composite null. We include the Sobel’s test, the MaxP test and the MT-Comp test (Huang 2019) for comparison. The exposure variable *A* was simulated from a Bernoulli distribution with success probability equal to 0.5. We simulated two continuous covariates *X*_1_ and *X*_2_ from *N* (10, 1) and *N* (5, 1) respectively, then the mediator *M* and the outcome *Y* were simulated as follows

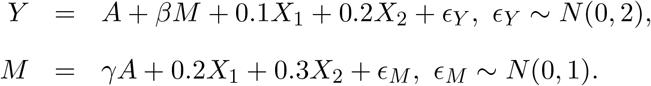

We first considered the following three extreme scenarios: (1) *w*_1_ = 1, *w*_2_ = 0, *w*_3_ = 0 where (*β, γ*) = (0.2, 0) ; (2) *w*_1_ = 0, *w*_2_ = 1, *w*_3_ = 0 where (*β, γ*) = (0, 0.2); (3) *w*_1_ = 0, *w*_2_ = 0, *w*_3_ = 1 where (*β, γ*) = (0, 0). The number of DNA methylation sites *m* was set to be 100,000. The significance levels *α* were: 0.05 and 0.01. Three sample sizes were considered: *N* = 500, 1000, 2000. In each scenario, we obtained *m p*-values for the exposure-mediator associations *p*_*γ,j*_, and *m p*-values for the mediator-outcome associations *p*_*β,j*_, where *j* = 1, 2, *· · ·, m*. We then applied all the testing procedures to calculate their empirical type I error rates. For the DACT method, we estimated the relative proportions of the three null cases based on these *m p*-value pairs and then applied the proposed DACT test. The type I error rates were then estimated as the proportions of *p*-values for mediation effect that are smaller than the significance levels *α*. The simulation results for those three scenarios are presented in Table 1.

**Table 1:** Empirical type I error rates of the four tests: Sobel’s test, MaxP test, MT-Comp and our DACT method under three nulls where (β, γ) are: (0.2, 0), (0, 0.2), (0, 0). The sample sizes are: 500, 1000 and 2000. The significance levels α are: 0.05, 0.01. The number of CpG sites m = 100, 000.

| Level $\alpha$ | $\beta$ | $\gamma$ | Sobel | | MaxP | | MT-Comp | | DACT | |
| --- | --- | --- | --- | --- | --- | --- | --- | --- | --- | --- |
|  |  |  | 0.05 | 0.01 | 0.05 | 0.01 | 0.05 | 0.01 | 0.05 | 0.01 |
| N=500 | 0.2 | 0 | 0.005 | 0.000 | 0.030 | 0.004 | 0.302 | 0.128 | 0.050 | 0.009 |
|  | 0 | 0.2 | 0.005 | 0.000 | 0.031 | 0.004 | 0.306 | 0.131 | 0.051 | 0.010 |
|  | 0 | 0 | 0.000 | 0.000 | 0.003 | 0.000 | 0.050 | 0.010 | 0.050 | 0.010 |
| N=1000 | 0.2 | 0 | 0.014 | 0.000 | 0.045 | 0.007 | 0.458 | 0.245 | 0.051 | 0.010 |
|  | 0 | 0.2 | 0.014 | 0.000 | 0.045 | 0.007 | 0.455 | 0.244 | 0.051 | 0.010 |
|  | 0 | 0 | 0.000 | 0.000 | 0.003 | 0.000 | 0.050 | 0.011 | 0.050 | 0.010 |
| N=2000 | 0.2 | 0 | 0.027 | 0.002 | 0.049 | 0.010 | 0.607 | 0.405 | 0.050 | 0.010 |
|  | 0 | 0.2 | 0.028 | 0.002 | 0.050 | 0.010 | 0.608 | 0.402 | 0.050 | 0.010 |
|  | 0 | 0 | 0.000 | 0.000 | 0.002 | 0.000 | 0.050 | 0.010 | 0.051 | 0.010 |

As shown in Table 1, the type I error rates of the Sobel’s test are smaller than the nominal significance levels in all three scenarios, especially in scenario 3. The type I error rates of the MaxP test get closer to the nominal significance levels in scenario 1 and 2 as the sample size increases. In scenario 3, increasing sample size does not change the empirical size of the MaxP test. The type I error rates of the MT-Comp method are inflated in scenario 1 and 2, and this inflation gets worse when the sample size increases. Huang (2019) also found that the MT-Comp can control type I error rate when the sample size is 500 or smaller. The MT-Comp method has correct size under scenario 3 and thus can work when the mediation effect signals are sparse with small sample sizes. The type I error rates of the proposed DACT are very close to the nominal levels in all three scenarios.

We next considered another three settings to assess the performance of the proposed DACT method mimicking real epigeome-wide mediation studies, where the three null cases are present for a fraction of DNA methylation sites. The total number of candidate mediators was *m* = 300, 000. We varied the relative proportions of *w*_1_, *w*_2_ and *w*_3_ = 1 −*w*_1_ −*w*_2_ to assess the performance of our method. Setting 1 (*w*_1_ = 0.33, *w*_2_ = 0.33, *w*_3_ = 0.34) represents the scenarios where the three cases are equally likely across the genome. Setting 2 (*w*_1_ = 0.05, *w*_2_ = 0.05, *w*_3_ = 0.90) represents the scenarios where Case 3 largely dominates. Setting 3 (*w*_1_ = 0.01, *w*_2_ = 0.01, *w*_3_ = 0.98) represents the scenarios that are often encountered in genome-wide epigenetic studies where Cases 1 and 2 are rare. Even in setting 3, there are 3000 mediators associated with the exposure only, and another set of 3000 mediators associated with the outcome only. In a typical epigenome-wide association study, the number of association signals is even smaller. We aim to demonstrate that our method can perform robustly even in those unfavorable settings. We simulate 300, 000 *Z*-test statistics (*Z*_*βj*_, *Z*_*γj*_) where *j* = 1, …, 300000. In Case 1, simulate *Z*_*βj*_ from *N* (*µ*_*β*_, 1) where *µ*_*β*_ is drawn from *N* (2, 1) and simulate *Z*_*γj*_ from *N* (0, 1). In Case 2, simulate *Z*_*βj*_ from *N* (0, 1) and *Z*_*γj*_ from *N* (*µ*_*γ*_, 1) where *µ*_*γ*_ is drawn from *N* (2, 1). In Case 3, simulate *Z*_*βj*_ from *N* (0, 1) and *Z*_*γj*_ from *N* (0, 1).

The QQ (quantile-quantile) plots for the *p*-values from uncorrected and corrected DACT using the estimated empirical null distribution are summarized in Figure 4. In setting 1, the uncorrected DACT is conservative while the corrected DACT works well. In setting 2, there is anoticeable difference between the uncorrected and corrected DACT methods. In setting 3, there is no noticeable difference between the corrected and uncorrected DACT method because the DACT statistic approximately follows uniform distribution on [0, 1], and thus the correction is usually not needed in such settings.

**Figure 4:**
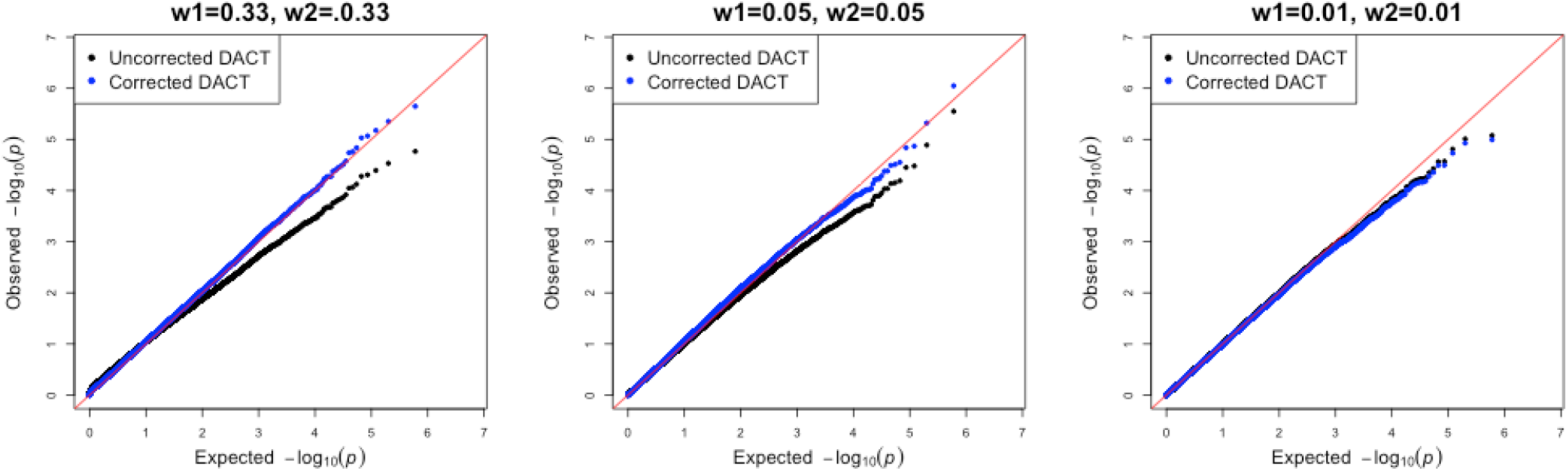
The QQ plots of the *p*-values for the uncorrected and corrected DACT method in three settings where *m* = 300, 000. The left-most figure is the QQ plot of uncorrected and corrected DACT for the setting where *w*_1_ = *w*_2_ = 0.33; the middle and right-most figures are the QQ plots where *w*_1_ = *w*_2_ = 0.05 and *w*_1_ = *w*_2_ = 0.01 respectively.

### 5.2 Power Comparison

Since the MT-Comp method has inflated type I error rates in Case 1 and Case 2, we do not include it for power comparison. The original Sobel and MaxP tests have deflated type I error rates and thus underpowered. At the significance level *α*, the power of Sobel’s test is estimated as the proportion of tests with *p*_*Sobel*_ *< α*, where *p*_*Sobel*_ is calculated using the standard normal approximation; the power of the MaxP test is estimated as the proportions of tests with *MaxP < α*. These two tests will serve as benchmarks for power comparison with the proposed DACT method.

We used the same simulation setup as that described in the first paragraph of Section 5.1 except that we simulated data under the alternative hypothesis. Specifically, we set (*β, γ*) to be the following values: (0.133, 0.3), (0.2, 0.2), (−0.3, 0.133) respectively, where the mediation effect size was set to be 0.04. The sample size *N* was set to be 500, 1000, or 2000. The number of DNA methylation sites *m* was 10, 000. The power was estimated as the proportion of rejections among those *m* tests at the significance level 0.05. The results are summarized in Table 2. As expected, the MaxP test is more powerful than Sobel’s test and the DACT test is more powerful than the MaxP test.

**Table 2:** Power comparisons of the Sobel’s test, MaxP test and the DACT test using simulation studies. The sample sizes considered are 800, 1000, 1200. The A − M and M − Y association effects (γ, β) are set to be (0.133, 0.3), (0.2, 0.2) and (−0.3, 0.133) where |γβ| = 0.04 in those three settings.

| $(\gamma, \beta)$ | (0.133, 0.3) | | | (0.2, 0.2) | | | (-0.3, 0.133) | | |
| --- | --- | --- | --- | --- | --- | --- | --- | --- | --- |
| $N$ | 800 | 1000 | 1200 | 800 | 1000 | 1200 | 800 | 1000 | 1200 |
| Sobel | 0.34 | 0.45 | 0.55 | 0.42 | 0.60 | 0.74 | 0.34 | 0.46 | 0.56 |
| MaxP | 0.46 | 0.55 | 0.62 | 0.65 | 0.78 | 0.87 | 0.45 | 0.55 | 0.63 |
| DACT | 0.47 | 0.55 | 0.63 | 0.76 | 0.87 | 0.93 | 0.46 | 0.56 | 0.64 |

We found that the power advantage of the DACT test over the MaxP test gets smaller with increasing differences in the magnitudes between *β* and *γ*. To investigate this matter, we further performed the following additional simulation studies. First, we set the mediation effect size *βγ* to be 0.04. Second, we divided the interval [0.04, 0.5] equally into 400 subintervals specified by 401 grid points *γ*_*j*_, and set *β*_*j*_ = 0.04*/γ*_*j*_ where *j* = 1, *· · ·*, 401. Under each alternative (*β*_*j*_, *γ*_*j*_), we performed one million simulations to estimate the powers of Sobel’s test, MaxP and DACT. We plotted the power estimates for all 401 grid points for the three tests: Sobel, MaxP and DACT. Figure 5 shows that the powers of the three tests are not monotone increasing functions of the mediation effect size *βγ*, but actually depend on the relative effect sizes of *β* and *γ*. The powers of these three tests are all maximized when |*γ/β*| = 1 and decrease quickly as |*γ/β*| deviates away from one. In other words, the powers of Sobel, MaxP and DACT are dictated by the smaller association signal of the *A* − *M* and *M* − *Y* associations. Those simulation results are in line with our theoretical findings in Results 1 and 2.

**Figure 5:**
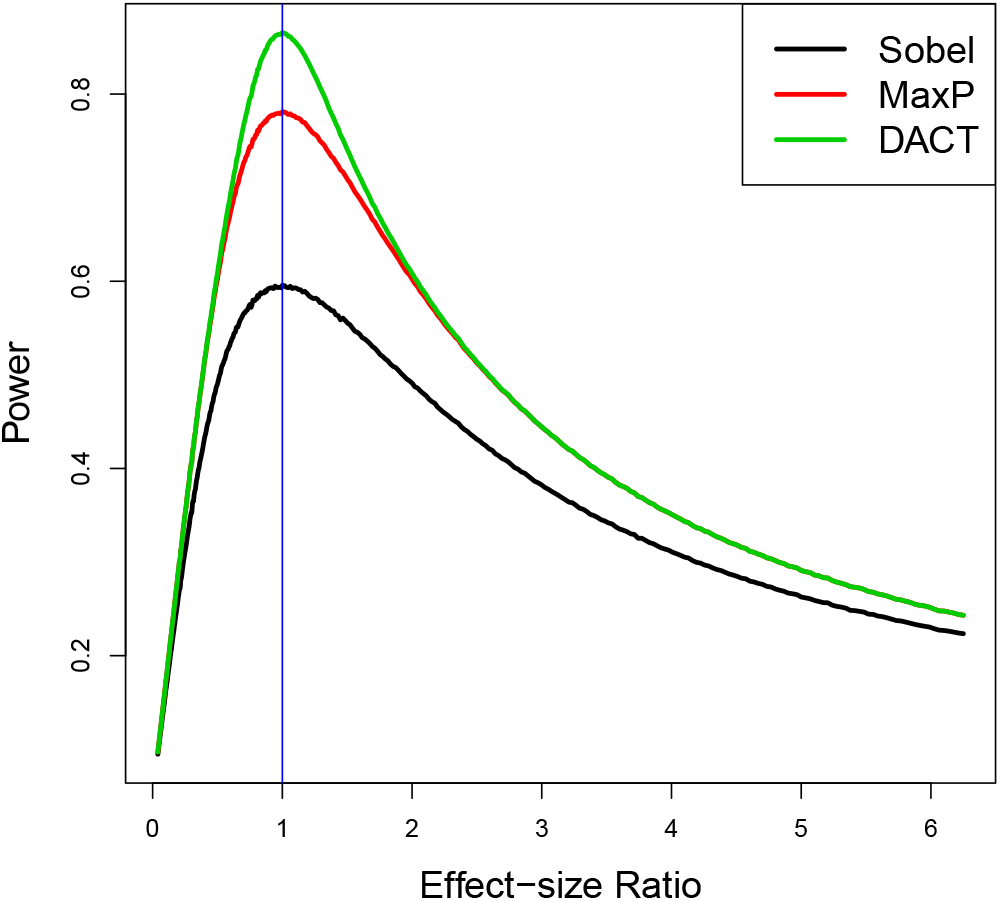
Power Comparison of the three tests: Sobel, MaxP and DACT using simulation studies. The same mediation effect size is fixed at 0.04 with different β and γ value combinations. The horizontal axis represents the effect size ratio |γ/β|.

To account for the correlation structure among DNA methylation sites, we performed an additional simulation study by simulating outcomes using the observed DNA methylation M-values of 24,264 CpG sites on chromosome 5 from the NAS data set (See Section 6 for more detailed background information), because we found strong mediation effect signals on this chromosome. In this numerical experiment, without loss of generality, we did not include covariates for simplicity. We set the sample size to be 603, the same as in the NAS data. We generated an exposure variable *A* from a Bernoulli distribution with probability 0.5. We then shifted the mean value of a randomly selected set of 2000 CpG sites among the exposed group (*A* = 1), and simulated the mean shift effect sizes from a uniform distribution on [−0.6, −0.2] mimicking the effect sizes of smoking on the methylation in the NAS data. We generated 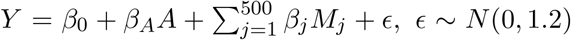, *ϵ* ∼ *N* (0, 1.2), where those 500 CpG sites were selected based on the most significant associations with the lung function from the analysis of the real NAS data, and the true coefficients *β*_*j*_, *j* = 1, …, 500, were set to be the estimated values from the NAS data. In this set-up, the numbers of CpG sites in the three null cases and the alternative case in (9) were: 500, 2000, 21723 and 41 respectively.

We repeated this numerical experiment 1000 times and estimated the FDR, and the mean true positive rate (TPP, or average power) (Dudoit and van der Laan 2007), which is defined as the proportion of mediation signals detected using the FDR threshold at 0.05. We included the MaxP test for comparison as the Sobel’s test has been shown to be less powerful than the MaxP test. For the MaxP test and the DACT method, we found that the estimated FDR was 0.042 (DACT) and 0 (MaxP), and the average power using FDR threshold is 0.86 (DACT) and 0.28 (MaxP). Therefore, the MaxP test was overly conservative, and the DACT method had an improved power while controlling for FDR at the nominal level in multiple testing settings.

## 6 The Normative Aging Genome-Wide Epigenetic Study

Cigarette smoking is an important risk factor for lung diseases (Anthonisen et al. 2002). Smoking behavior has been found to be associated with DNA methylation levels (Breitling et al. 2011; Li et al. 2018), and DNA methylation levels have also been found to be associated with lung functions (Lepeule et al. 2012). It is thus of scientific interest to identify DNA methylation CpG sites that may mediate the effects of smoking on lung functions. Previous research has found two CpG sites (cg05575921, cg24859433) as mediators lying in the causal pathway from smoking to lung functions using underpowered testing procedures (Zhang et al. 2016; Barfield et al. 2017). In this section, we demonstrate that the proposed DACT method has improved power to detect more DNA methylation CpG sites that might mediate the effect of smoking on lung functions.

The Normative Aging Study (NAS) is a prospective cohort study established in Eastern Massachusetts in 1963 by the U.S. Department of Veteran Affairs (Bell et al. 1972). The men were free of known chronic medical conditions at enrollment, and returned for on-sites, follow-up visits every 3-5 years. During these visits, detailed physical examinations were performed, bio-specimens including blood were obtained, and questionnaire data pertaining to diet, smoking status, and additional lifestyle factors that may impact health were collected. The DNA methylation was measured using the Illumina Infinium HumanMethylation450 Beadchip on blood samples collected after an overnight fast (Bibikova et al. 2011). After quality control, methylation Beta-values ranging from 0 (no methylation) to 1 (full methylation) was calculated for each CpG site (Teschendorff et al. 2012). We then use the logit (base 2) function to transform the Beta-values into M-values for statistical analysis because the M-value scale is more statistically valid for regression models as it is approximately homoscedastic (Du et al. 2010). The batch effects were adjusted by the ComBat algorithm (Johnson et al. 2007). In total, we had DNA methylations measured for 484,613 CpG sites on 603 men.

The binary exposure was smoking status (current or former smokers versus never smokers), and the outcome was the forced expiratory flow at 25%-75% of the Forced Expiratory Vital capacity (FEF_25−75%_). We transformed FEF_25−75%_ using squared root to achieve better normality. We adjusted for age, height, weight, education history, medication history, blood cell type abundances (Houseman et al. 2012), and five principal components (previously calculated to represent 95% of DNA processing batch effects), all based on our prior work studying DNA methylation in this cohort. We then fit the outcome and mediator linear regression models and obtain *p*-values for *γ* (smoking - methylation) and *β* (methylation - lung function) for each of the 484,613 CpG sites (the QQ plots are given in Figure S1 in the Supplementary Materials). The proportions of nulls for the parameter *γ* and *β* were estimated as 0.996, 0.9867 respectively using the JC method (Jin and Cai 2007). Using equation (9), the proportions of the four cases were estimated as (0.01423, 0.00416, 0.98155, 0.0006). Therefore, the mediation effect signals were very sparse in the NAS data set.

Under the composite null, the relative proportions of the three null cases (after normalization) were estimated as *ŵ* _1_ = 0.014, *ŵ* _2_ = 0.004, *ŵ* _3_ = 0.982. We then computed the DACT, performed inverse normal CDF transformation to obtain *z*-scores. A histogram of the transformed DACT (*z*-scores) indicates strong normality as shown in Figure 6 (the left sub-figure). The mean *δ* and the standard deviation *σ* of the null distribution *N* (*δ, σ*^2^) were estimated as 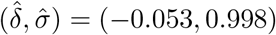 using equation (15) in Section 3.2.

**Figure 6:**
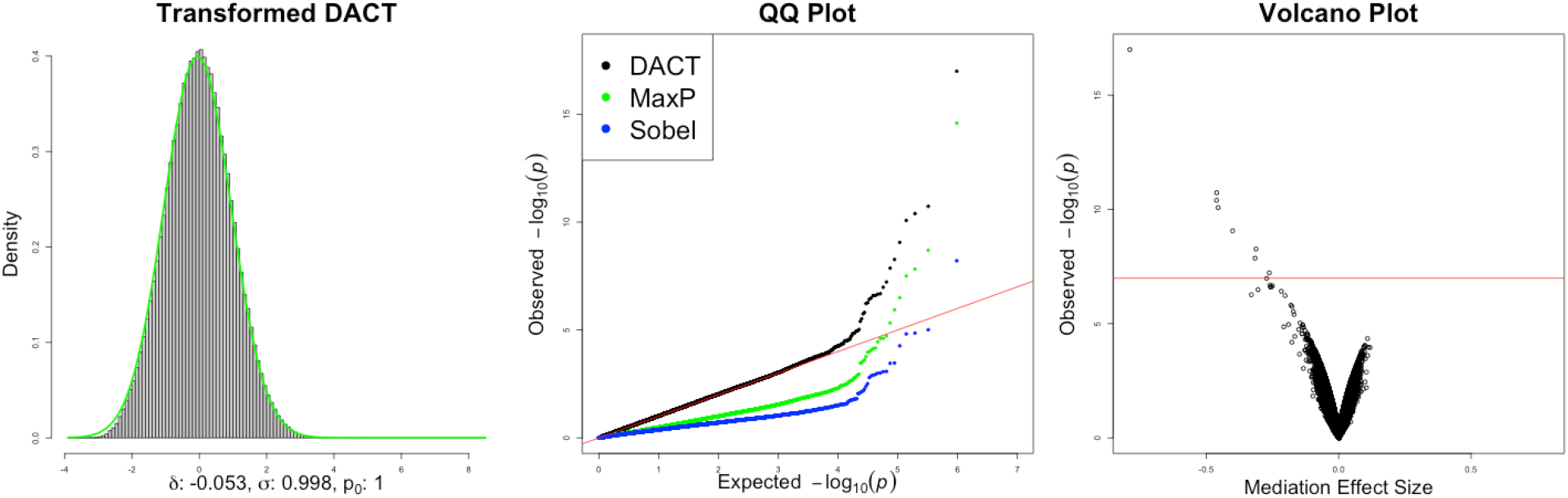
The left sub-figure is a histogram of the z-scores transformed from the DACT statistics based on the inverse normal cumulative distribution function. The green solid line is the estimated empirical null density function with mean −0.053 and standard deviation 0.998 using equation (15). The middle one is the QQ plot of the Sobel’s test, the MaxP test and the corrected DACT method. The right one is the volcano plot for the corrected DACT method, where the horizontal axis represents the mediation effect sizes and the vertical axis represents the corrected p-values of the DACT method on the − log_10_ scale.

The QQ plot in Figure 6 (the middle sub-figure) showed that both Sobel’s test and the MaxP test produced seriously deflated *p*-values and hence were underpowered to detect CpG sites with mediation effects. In contrast, the proposed DACT method performed very well, and its genomic inflation factor was estimated as *λ* = 1.07. The volcano plot in Figure 6 (the right sub-figure) showed that those more significant CpG sites also tended to have larger mediation effect sizes, and thus the statistical significance was mainly driven by the large effect sizes rather than small standard errors.

Using the tail FDR threshold at 0.05, we found 19 mediation effect signals summarized in Table S1 in the Supplementary Materials. To save space, we present the most significant top eight CpG sites in Table 3. Those CpG sites are also significant using the more stringent Bonferroni corrected threshold (0.05*/*484613 = 1.03 *×* 10^−7^). A Manhattan plot is also provided in Figure S2 in the Supplementary Materials. In Table 3, the Sobel’s test only detected CpG site cg05575921, and the MaxP test detected four CpG sites: cg05575921, cg03636183, cg06126421 and cg21566642. The proposed DACT method further detected additional CpG sites that were missed by the Sobel’s test and the MaxP test.

**Table 3:**
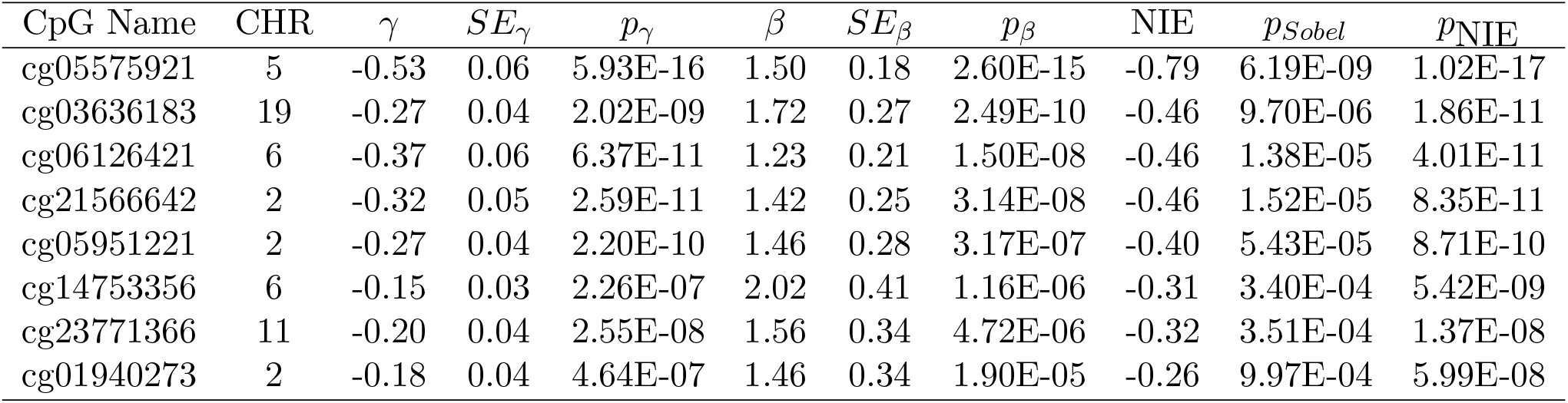
Top hits from causal mediation analysis of the Normative Aging Genome-wide Epigenetic Study. The exposure is smoking status, and the outcome is lung function measure FEF_25− 75%_. CHR stands for chromosome number. NIE stands for Natural Indirect Effect (mediation effect). The p_NIE_ column is computed using the DACT method after correction.

| CpG Name | CHR | $\gamma$ | $SE_{\gamma}$ | $p_{\gamma}$ | $\beta$ | $SE_{\beta}$ | $p_{\beta}$ | NIE | $p_{Sobel}$ | $p_{NIE}$ |
| --- | --- | --- | --- | --- | --- | --- | --- | --- | --- | --- |
| cg05575921 | 5 | -0.53 | 0.06 | 5.93E-16 | 1.50 | 0.18 | 2.60E-15 | -0.79 | 6.19E-09 | 1.02E-17 |
| cg03636183 | 19 | -0.27 | 0.04 | 2.02E-09 | 1.72 | 0.27 | 2.49E-10 | -0.46 | 9.70E-06 | 1.86E-11 |
| cg06126421 | 6 | -0.37 | 0.06 | 6.37E-11 | 1.23 | 0.21 | 1.50E-08 | -0.46 | 1.38E-05 | 4.01E-11 |
| cg21566642 | 2 | -0.32 | 0.05 | 2.59E-11 | 1.42 | 0.25 | 3.14E-08 | -0.46 | 1.52E-05 | 8.35E-11 |
| cg05951221 | 2 | -0.27 | 0.04 | 2.20E-10 | 1.46 | 0.28 | 3.17E-07 | -0.40 | 5.43E-05 | 8.71E-10 |
| cg14753356 | 6 | -0.15 | 0.03 | 2.26E-07 | 2.02 | 0.41 | 1.16E-06 | -0.31 | 3.40E-04 | 5.42E-09 |
| cg23771366 | 11 | -0.20 | 0.04 | 2.55E-08 | 1.56 | 0.34 | 4.72E-06 | -0.32 | 3.51E-04 | 1.37E-08 |
| cg01940273 | 2 | -0.18 | 0.04 | 4.64E-07 | 1.46 | 0.34 | 1.90E-05 | -0.26 | 9.97E-04 | 5.99E-08 |

The top CpG site cg05575921 is located in the aryl-hydrocarbon receptor repressor (AHRR) gene on chromosome 5 and has been consistently found to be demethylated among smokers compared to non-smokers (Joubert et al. 2012; Philibert et al. 2012, 2013; Reynolds et al. 2015). It has also been found to be associated with increased lung cancer risk (Fasanelli et al. 2015). Previous mediation analysis using the underpowered MaxP test can also detect this CpG site cg05575921 as an mediator in the pathway from smoking to lung functions (Zhang et al. 2016; Barfield et al. 2017), simply because the *p*-values for the smoking-methylation and methylation-lung functions associations were both highly significant.

The CpG site cg03636183 in F2RL3 was also found to be a biomarker of smoking exposure (Zhang et al. 2014) and was related to mortality among patients with stable coronary heart disease (Breitling et al. 2012) and increased lung cancer risk (Fasanelli et al. 2015). It has been found that the CpG site cg06126421 in the intergenic region at 6p21.33 to be hypomethylated among smokers compared to non-smokers (Shenker et al. 2012; Elliott et al. 2014). The CpG site cg06126421 was found to be associated with all-cause, cardiovascular, and cancer mortality, for participants with methylation levels in the lowest quartile of this CpG site (Zhang et al. 2016). The CpG sites cg21566642 and cg05951221 located on the same CpG island of chromosome 2 were found to be associated with increased lung cancer risk (Fasanelli et al. 2015). Our analysis suggests that those significant CpG sites might play important biological roles in mediating the effect of smoking on lung functions.

To check for any possible violation of the no unmeasured confounding assumption, we further performed a comprehensive sensitivity analysis to assess the robustness of our mediation analysis results to any unmeasured confounding variables. The idea is that the residual correlation *ρ* between the two error terms in the linear mediator and outcome regressions are correlated if the unmeasured confounding assumption is violated and vice versa (Imai et al. 2010). Therefore, the residual correlation *ρ* can be used to measure the magnitude of confounding bias, where *ρ* = 0 implies no confounding bias. We can hypothetically vary *ρ* to observe the change to the mediation effect estimates. When |*ρ*| deviates from zero to some extent, the observed mediation effects could be explained away by the confounding bias. We varied the value of *ρ* and computed the corresponding value of NIE using the R package *mediation* (Tingley et al. 2013). We found that to explain away the mediation effects of CpG sites cg05575921 and cg03636183 in the causal pathway from smoking to lung function, the confounding bias measured by *ρ* needs to be at least 0.3, and to explain away the mediation effects of the other CpG sites provided in Table 3, *ρ* needs to be at least 0.2. Such large confounding bias is absent in our data analysis, as we found that the residual correlation *ρ* for all the eight CpG sites are very close to zero with absolute value smaller than 10^−17^, showing that the confound bias is negligible. Our sensitivity analysis results show that we have adjusted sufficient covariates in the mediation analysis for all the CpG sites in Table 3. Therefore, our mediation analysis results are robust to unmeasured confounding. More detailed sensitivity analysis results are provided in the Supplementary Materials.

## 7 Discussion

In this paper, we developed a valid and powerful testing procedure for detecting DNA methylation CpG sites that might mediate the effect of an exposure on an outcome of interest in genomewide epigenetic studies. Despite that the Wald-type Sobel’s test and the likelihood ratio test equivalent MaxP test were empirically found to have low power for decades, however, no successful remedy has been proposed to resolve the conservativeness of the two tests. A lack of method development for this problem is incompatible with the increasing need of powerful testing procedures for detecting mediation effects in large-scale epigenetic studies. Testing a large number of composite nulls leverages the two sides of the same coin. On one side, multiple testing correction is a curse and makes it more challenging for the inference of mediation effects than the single mediation effect testing problems. But on the other side, multiple testing for mediation effects is a blessing because it enables us to estimate the relative proportions of the three null cases that can be leveraged to improve power.

Understanding the reasons why Sobel’s test and the MaxP test are conservative paves the way for developing a more powerful test. We found that the null Case 3 is the singular point in the null parameter space, under which the standard asymptotic argument fails. We show that the MaxP test is essentially the likelihood ratio test for the composite null of no mediation effect, but it does not follow the traditional chi-squared distribution with one degree of freedom (on the *Z*^2^ scale) but rather follows Beta distribution *Beta*(2, 1) in the null Case 3. The Wald-type Sobel’s test does not follow the standard normal distribution in the null Case 3 either, instead it follows the normal distribution with mean zero and variance equal to one quarter which can be shown by the not so well-known “super Cauchy phenomenon” (Pillai and Meng 2016). Those important discoveries provide rigorous explanations on why the widely used Sobel’s test and the MaxP test are underpowered for inferring the presence of mediation effects in both single test and multiple testing scenarios, more importantly, inspire us to develop the DACT method.

Our contributions are multi-fold. First, we divide the null parameter space into three disjoint parts and find that the null Case 3 is the culprit of the poor performances of Sobel’s test and the MaxP test. Such a decomposition also inspires us to obtain correct case-specific *p*-values. Second, we leverage the genome-wide data to consistently estimate the relative proportions of the three null cases and then construct the DACT test, turning the curse of multiple testing into a blessing. Third, large-scale testing also permits the use of empirical null distribution for inference. This approach is especially useful when the exposure-mediator or/and mediator-outcome association signals are non-sparse. Fourth, the DACT procedure is computationally fast and is scalable for large-scale inference of mediation effects. We also developed an user-friendly R package *DACT* for public use. Our NAS data analysis findings are of scientific interests. Detection of DNA methylation CpG sites that may mediate the effect of smoking behavior on lung function can help us understand the underlying causal mechanism and biological pathway of the observed association between smoking and lung function. These identified CpG sites can also be used as intervention targets to reduce the harmful effects of smoking on lung function. Previously, only two CpG sites with strong signals have been found as putative mediators in the causal pathway from smoking to lung function (Barfield et al. 2017). A lack of powerful tests hindered researchers to discover more potential mediators. We applied the newly developed DACT procedure to the Normative Aging Study and identified additional DNA methylation CpG sites that were missed by previous analysis. Our comprehensive sensitivity analysis suggests that the mediation analysis results are robust to unmeasured confounding factors.

The proposed DACT procedure is developed for genome-wide epigenetic studies where we can estimate the relative proportions of the three cases under the composite null hypothesis. Notice that accurate estimation of these proportions is crucial for performing the DACT test, especially when the *p*-values across the CpG sites are correlated. The JC method for estimating these proportions was found to be accurate and consistent in both sparse and non-sparse settings even for dependent data, and has been adopted in our DACT procedure. It is of future research interest to extend the DACT method to the setting in which there are a large number of exposures, e.g, genetic variants in Genome-Wide Association Studies, as well as univariate or multivariate mediators. When the binary outcome is not rare, the NIE is no longer equal to *βγ* even approximately (Gaynor et al. 2019). Testing NIE in those settings is challenging and is of future research direction. Our DACT procedure is not applicable for a single mediation test if the relative proportions of the three null cases cannot be empirically estimated. It is hence of future research interest to develop powerful mediation tests in such settings.

## Software

The DACT procedure was implemented in the R package *DACT*, which is publicly available at https://github.com/zhonghualiu/DACT.

## Data Availability

Individual level data is not publicly available.

## Supplementary Materials

The online supplementary materials provide technical proofs and additional data analysis results.

## Notes

* This work was supported by the National Institutes of Health grants R35-CA197449, P01-CA134294, U01-HG009088, U19-CA203654, R01-HL113338, P30 ES000002 and T32GM074897. This work was also supported by Dr. Zhonghua Liu’s start-up research fund (000250348) from the University of Hong Kong. We would like to thank the editors and reviewers for their helpful comments that improved the paper.

### Competing Interest Statement

The authors have declared no competing interest.

### Funding Statement

This work was supported by the National Institutes of Health grants R35-CA197449, P01-CA134294, U01-HG009088, U19-CA203654, R01-HL113338, P30 ES000002, and T32GM074897. This work was also supported by Dr. Zhonghua Liu's start-up research fund (000250348) from the University of Hong Kong.

### Author Declarations

All participants provided written informed consent to the VA Institutional Review Board (IRB), and both the Harvard T.H. Chan School of Public Health and VA IRBs granted human subjects approval.

### Summary of Updates

Revised abstracts.

